# Predictors of mortality among COVID-19 patients at Kilimanjaro Christian Medical Centre in Northern Tanzania: A hospital-based retrospective cohort study

**DOI:** 10.1101/2024.03.05.24303842

**Authors:** Gilbert G. Waria, Florida J. Muro, Norman Jonas, Francis Sakita, Laura Shirima, Henry L. Mlay, Michael Ngowi, Elvis B. Meela, Innocent B. Mboya, Kajiru G. Kilonzo

**Affiliations:** Department of Epidemiology and Biostatistics, Institute of Public Health, Kilimanjaro Christian Medical University College, Moshi Tanzania; Department of Community Medicine, Institute of Public Health, Kilimanjaro Christian Medical University College, Moshi, Tanzania; Kilimanjaro Christian Medical Centre, Moshi, Tanzania; Department of Internal Medicine, Kilimanjaro Christian Medical University College, Moshi, Tanzania; Department of Translational Medicine, Lund University, Malmö, Sweden; Emergency Medicine Department, Kilimanjaro Christian Medical Centre, Moshi, Tanzania

**Keywords:** Mortality rate, Mortality, Predictors, COVID-19, Tanzania

## Abstract

**Background:** COVID-19 disease is a global public health disaster causing a range of social, economic, and healthcare difficulties, border restrictions, high human loss, lockdown, and transportation challenges. Despite it being a global pandemic, there are few studies conducted in Tanzania to examine the predictors of mortality. This disease has caused a significant number of mortalities worldwide but literature shows low mortality and better survival in Africa than in other WHO regions. Therefore, this study aimed to determine the predictors of mortality among COVID-19 patients at KCMC Hospital in Northern Tanzania.

**Methodology:** This was the hospital-based retrospective cohort study, conducted at KCMC Hospital in Northern Tanzania among all admitted patients with confirmed COVID-19, from 10^th^ March 2020 to 26^th^ January 2022. The main study event was COVID-19 mortality. The predictors of mortality were determined by using the Weibull survival regression model and the statistically significant results were declared at a p-value of <0.05.

**Results:** A total of 547 confirmed COVID-19 patient records were included in the study. Their median age was 63 (IQR; 53-83), about 60% were aged 60 years and above, and 56.7% were males. The most common clinical features were; fever (60.8%), a severe form of the disease (44.4%), difficulty in breathing (73.3%), chest pain (46.1%), and generalized body weakness (71.3%). Of all participants, over one-third (34.6%) died (95%CI; 0.31-0.39). The median survival time was 7 days (IQR; 3-12). The overall mortality rate was 32.33 per 1000 person-days while the independent predictors of higher mortality risk were age ≥60 years (AHR=2.01; 95%CI 1.41-2.87; P<0.001), disease severity (AHR=4.44; 95%CI 2.56-7.73; P<0.001) and male sex (AHR=1.28; 95%CI; 0.93-1.73; P=0.128).

**Conclusion:** Mortality was higher in elderly male patients, with a severe form of the disease and those with any comorbidities. Therefore, more attention should be provided to older patients including uptake of the current vaccine and ensuring standard and supportive care at primary health facilities is available.

## Introduction

On 31 December 2019, the World Health Organization (WHO) was informed of a cluster of cases of pneumonia of unknown cause detected in Wuhan, Hubei Province, China. This condition is due to the severe acute respiratory syndrome coronavirus-2 (SARS-CoV-2), causing a range of clinical signs including fever, cough, asthenia, or respiratory distress. The virus has spread rapidly worldwide since its outbreak with the WHO declaring it a pandemic [1] As of July 8, 2022, more than 500 million cases of COVID-19 have been reported across the world, resulting in more than 6 million deaths[2].

The global COVID-19 data showed America, Asia, and Europe to be the most hit continents with COVID-19 with poor survival rates among cases as compared with other continents like Africa which had better survival [1]. The first confirmed case of COVID-19 in Africa was reported in Egypt on February 14, 2020, and on April 19, 2020, Egypt has reported 3144 cases of COVID-19 and 239 deaths. The case fatality rate of COVID-19 varies across countries with an average of 4.2 ± 3.8, among the older population [4, 5]

In China, Most of the mortality cases were reported among men, and less than half had underlying diseases including diabetes, hypertension, and cardiovascular disease [6, 7]. There is a better survival rate for COVID-19 patients, who are not living with comorbidities, such as hypertension, diabetes, liver disease, renal disease, neurological disease, and immunocompromised [8].

In 2020, South Africa, Egypt, Morocco, Nigeria, and Ghana were the African countries with the highest number of cases of COVID-19 [9]. Mortality was higher among older individuals (60+ years), with chronic disease and travel history, but with a lower mortality rate of 5.6% compared to European countries [10]. Nevertheless, Africa has been threatened by insufficient infrastructure and weak healthcare systems, putting the social-economic status of Africa under control [11]. Some highlights in Africa that have been mentioned as predictors of low mortality include high temperature, humidity, early use of BCG vaccination, and possibly a different viral subtype [12].

In Tanzania, the first case of COVID-19 was imported on 16^th^ March 2020 and declared by the Ministry of Health, Community Development, Gender, Elderly, and Children [MoHCDGEC] on 14^th^ April 2020 that there was a community transmission, with more than five hundred (509) confirmed cases and 21 deaths related to COVID 19 [13]. Currently, we have more than 35,000 confirmed cases and more than 800 deaths as of July 8, 2022 [14]

Interventions to improve survival among COVID-19 cases include ongoing vaccination, Infection Prevention and Control (IPC) policy through the use of personal protective equipment/gear, continuous education, community engagement, and protection of high-risk groups including healthcare workers, older adults, and those with underlying conditions or comorbidities [14–18]. However, the number of new infections and deaths attributed to COVID-19 continues to spike in various communities and regions. In 2021, Kilimanjaro and Mwanza regions which are surrounded by Kenya and Uganda in the Northern and Northwestern part of Tanzania were the most regions affected by the disease [14]Currently, more than 3 Million people have received at least one COVID-19 vaccine dose in Tanzania [19]. Despite that, Tanzania lacks studies documenting COVID-19-related mortality and associated predictors. Therefore, this study aimed to determine predictors of mortality among COVID-19 patients admitted at a zonal referral hospital in Northern Tanzania.

## Methods

### Study design, setting, and population

This was a hospital-based retrospective cohort study, which involved the analysis of all COVID-19 patients admitted at the KCMC hospital from 10^th^ March 2020 to 26^th^ January 2022 comprising data from all four COVID-19 waves in Tanzania. We used data available in Excel format of the electronic isolation database. The data contained the patient’s information on socio-demographic characteristics, clinical features, and comorbidities. The electronic patient records were collected and filled in by the clinicians at clinical examinations. A total of 1117 COVID-19 patient records were retrieved from the electronic files, only 570 Confirmed COVID-19 were enrolled. The study excluded patients with incomplete information such as the unknown discharge status, and dates of admission and discharge (n=23), which was crucial for our statistical analyses and after removing duplicate patient records only 547 confirmed COVID-19 cases were available for analysis **(S1 Fig)**.

### Variables

The main event of interest was COVID-19 mortality which was defined based on the certified cause of death due to COVID-19 documented in the patient’s electronic files. The earliest observed date was 10^th^ March 2020 while the latest exist date was 26^th^ Jan 2022.

Independent variables included patients’ socio-demographic characteristics, clinical features at admission, and comorbidities. The study’s selection and categorization of independent variables were based on previous literature [16]. Patients’ socio-demographic characteristics included age in years (<60, ≥60 years), sex (male, female), and residence region (Arusha, Kilimanjaro, others). Clinical features at admission were measured based on the defined WHO cutoff points; Disease severity, Mild case if Oxygen saturation ≥94% on RA+ No clinical or radiological evidence of pneumonia, Moderate case if oxygen saturation 90-94% on RA+ clinical or radiological evidence of non-severe pneumonia, Severe case if oxygen saturation 80-89% on RA+ clinical or radiological evidence of severe pneumonia. COVID-19 symptoms, specifically fever, headache, nausea, cough, difficulty in breathing, chest pain, generalized body weakness, sore throat, runny nose, smell loss, loss of taste, abdominal pain, and diarrhea were recorded and measured based on patient/relative report and clinician assessment (“Yes” if had any symptom and “No” if had no symptoms).

Furthermore, the comorbidities were the presence of hypertension, diabetes, liver disease, asthma, renal disease, neurological disease, immunodeficiency, and malignancy. Patients were also considered to have comorbidities if they had a history of comorbidities or taking any anti-comorbidities drugs such as antihypertensive, antidiabetic, antiviral, anticancer drugs, and being under current care/management receiving (Yes/No). Abnormal chest x-ray and abnormal lung auscultation were defined based on the radiological finding.

### Data analysis

We performed all statistical analyses using Stata version 15 (Stata Corp. 2017). Stata statistical software: Release 15. College Station, TX: StataCorp LLC). We summarized numerical variables (age and Oxygen saturation at admission) using median (interquartile range) and categorical variables using frequencies and percentages. The Chi-square test was used to test the association between mortality and socio-demographic characteristics, clinical features, and comorbidities at a 5% threshold level. We compared patient survival experiences using the Kaplan-Meier survival function and Nelson-Aalen cumulative hazard plots with the Log-rank test. The mortality rate was estimated using the Poisson survival regression model while the Weibull regression model was used to estimate the associated predictors [21]. Model comparison was done using the Akaike Information Criteria (AIC), the lowest value indicating the best-fitting model.

Crude/unadjusted Weibull regression determined the association between mortality and participant characteristics. The crude hazard ratio (CHR) and corresponding 95% confidence intervals (CI) were reported for variables of clinical importance such as hypertension, diabetes, age, and sex. We further developed a multivariable Weibull regression model to determine mortality predictors adjusted for potential confounders. The model estimated the adjusted hazard ratio (AHR) and the corresponding 95% CI.

### Ethical consideration

Permission to use the COVID-19 data with reference No.KCMC/P.I/Vol.IX was granted by the KCMC Hospital executive director’s office. Unique patient identification instead of hospital numbers was used to maintain confidentiality. Ethical approval to carry out the current study was obtained from the Kilimanjaro Christian Medical College Research Ethics and Review Committee (KCMU-CREC) with clearance number PG 15/2021.

## Results

### Characteristics of the studied COVID-19 patients

Among 547 patients who tested positive for COVID-19 and met inclusion criteria, 324 (60%) were aged 60 years and above with a median age of 63 (IQR: 50-75), and more than half of them were males 310 (56.7%) **(S1 Table).** Most of the study participants presented with fever 333 (60.8%), a severe form of COVID-19 243 (44.4%), difficulty in breathing 424 (73.3%), chest pain 252 (46.1%), and generalized body weakness 390 (71.3%). Two-thirds of the participants 375 (68.6%) were exposed during waves 2 & 3; their median oxygen saturation at the time of attending the hospital was 70% (IQR: 53-82) **(S2 Table)**. Abnormal chest x-ray 458 (83.7%), abnormal lung auscultation 274 (50.1%), hypertension 255 (46.6%), and diabetes 159 (29.1%) were the most common comorbidities **(S3 Table)**.

### Proportions of COVID-19 deaths by participant characteristics

Of 547 COVID-19 patients, 189 (34.6%) died and 358 (65.5%) were discharged alive. The median survival time was 7 days (IQR:3-12 days). The overall mortality rate was 32.33, 95%CI; 27.8-37.3 per 1000 person-days. Mortality occurred in 141 (43.5%) of the elderly patients (P<0.001) and 113 (36.5%) in male patients (P=0.285) **(S2 and S3 Table)**. The proportion of deaths was higher in 140 (57.6%) patients with a severe form of the disease (p<0.001), among 154 (38.4%) patients who present with difficulty in breathing (p=0.002), 133 (35.5%), 31 (43.7%) in patients exposed during waves 2&3 and 4, respectively (p=0.03), 19 (25.7%) who reported to loss of taste (p-value =0.084) and in 10 (22.7%) among those who reported having had diarrhea episodes (p-value=0.085) **(S2 Table)**.

Among individuals with comorbidities, 66 (41.5%) patients with diabetes (p=0.028), 172 (37.6%) with abnormal chest x-ray (p<0.001), 110 (40.2%) with abnormal lung auscultation (p=0.006), and 13 (54.2%) = with renal disease (p=0.003) had died **(S3 table)**.

### Mortality rate experience of COVID-19 patients

The overall mortality rate was 32.33, 95%CI; 27.8-37.3 per 1000 person-days. The mortality rate (MR) per 1000 person-days was 46.7 (95%CI; 39.4-55.4) in patients over 60 and (MR=37.1, 95%CI; 30.8-44.5) in male patients. Furthermore, patients with moderate and severe forms of the disease had mortality rates of (MR=19.2, 95%CI; 13.3-27.4) and (MR=55.0, 95%CI; 46.4-65.2), respectively. The mortality rate (per 1000 person-days) among patients presenting with breathing difficulties was (MR=35.4, 95%CI; 30.1-41.6), and in diabetic patients was (MR=39.4, 95%CI; 30.7-50.6). In addition, the mortality rate was (MR=36.9, 95%CI; 30.0-45.2) in hypertensive patients, (MR=34.5, 95%CI; 29.6-40.2) among patients with abnormal chest x-rays, and (MR=37.8, 95%CI; 31.2-45.9) among those with lung auscultations. Patients with renal disease had a mortality rate of (MR=37.8, 95%CI; 31.2-45.9) per 1000 person-days (**S2 Fig**).

### Predictors of COVID-19 mortality

The predictors associated with COVID-19 mortality are displayed in **(Tables S4, S5 & S6)**. In crude analysis Age, Sex, Disease severity, Difficult in breathing, Abnormal chest X-ray, Abnormal Lung auscultation and renal diseases were the significant predictor, the hazard of mortality among patients aged ≥60 years was 2.62 times significant higher, compared to those aged <60 years (CHR=2.62; 95%CI 1.88-3.66; P <0.001). Males had 1.35 times significant higher the hazard of mortality compared to females (CHR=1.35; 95%CI 1.00-1.83; P= 0.049). Compared to the mild form of the disease, patients with moderate form had a 1.91 times higher hazard of mortality (CHR=1.91; 95%CI 1.03-3.53; P <0.041). Compared to the mild form of the disease, patients with the severe form had a 5.54 times higher hazard of mortality (CHR=5.54; 95%CI 3.25-9.44; P <0.001). Patients who present with Difficult breathing had 1.56 times the significant higher hazard of mortality compared to those without (CHR=1.56; 95%CI 1.06-2.28; P=0.023), compared to patients living without diabetes, diabetes patients had 1.33 times the significant higher hazard of mortality (CHR=1.33; 95%CI 0.98-1.82; P= 0.066). Patients found with abnormal chest X-rays had 1.75 times the significant higher hazard of mortality compared to patients without abnormal chest x-ray, (CHR=1.75; 95%CI 1.06-2.88; P= 0.028). Patients with abnormal lung auscultation had 1.41 times the significant higher hazard of mortality compared to a patient with normal lung auscultations (CHR=1.41; 95%CI 1.05-1.89; P= 0.024). Lastly, compared to patients without renal disease, patients with renal disease had a 1.89 times higher hazard of mortality (CHR=1.89; 95%CI 1.08-3.33; P= 0.027).

In the adjusted analysis, the significantly higher mortality risk was among the elderly aged ≥60 years (AHR=2.01; 95%CI 1.41-2.87; P<0.001), male patients (AHR=1.27; 95%CI 0.93-1.73; P=0.128), patients diagnosed with moderate (AHR=1.67; 95%CI 0.89-3.13; P=0.112) and severe form of COVID-19 (AHR=4.44; 95%CI 2.56-7.73; P<0.001), those exposed during wave 2 & 3 (AHR=1.58; 95%CI 1.03-2.41; P=0.036), and in diabetes patients (AHR=1.25; 95%CI 0.88-1.79; P=0.212) **(S7 Table)**.

## Discussion

This study aimed to determine the predictors of mortality among COVID-19 patients at KCMC referral Hospital in northern Tanzania. The overall proportion of death due to COVID-19 among the studied patients was 34.6%, which is consistent with findings in Italy at 37.6% [3], Democratic republic of Congo at 32% [16], and in Dar es Salaam Tanzania at 31.8% [18]. Much lower prevalence was reported in India (14.4%) [19] and Tanzania (14.9%) [20], due to severe form of disease,, pre-existing comorbidities, and advanced age. The overall mortality rate in this study was 32.3 per 1000 person-days, which is lower than those reported in other settings [25–29]. Possible explanation may be the healthcare setting infrastuctures, Manpower and its accessibility, geographical location, or personal behaviors.

The median survival time was 7days (IQR:3-12] Similar to findings in East India 8days (IQR:7-10) [30], Ethiopia 9days (IQR:8-12), [26] USA 5days(IQR:3-10) [25],Tanzania 3days(IQR:1-6] [24], 12days Congo [16],Different findings in Ethiopia 44days(IQR:28-74) [29] Probable reasons living environment, Setting in which the patient receive care and attitudes toward COVID-19.

Older patients had a higher mortality risk compared to patients less than 60 years. The finding is similar to studies conducted in Northern Italy [5], Brazil [25], the Democratic Republic of Congo [16], and Tanzania [18]. This observation amongst the elderly may be due to immunodeficiency, and comorbidities, as it increases with age, which results in higher morbidity and mortality [26].

Mortality among males with COVID-19 was higher compared to females, which is similar to studies conducted elsewhere [20, 24, 32, 34–38]. The reasons for higher risk in males compared to females could be due to sex-specific hormones, and immune responses. Higher expression of Angiotensin-converting enzymes, Risk-taking behaviors and lifestyle, comorbidities, irresponsible attitudes toward the COVID-19 pandemic frequent hand washing, wearing masks and homestay, and hormonal difference [37, 38].

COVID-19 mortality was significantly higher among patients with moderate (18.3%) and severe forms of the disease (57.6%) compared to those with mild forms (13.2%). Patients with moderate and severe forms of the disease are consistently reported to have a higher mortality risk [23, 34, 39–42]. Weak immune functions, elderly age, limited organ compensatory function, more basic disease before infections, poor history of healthcare-seeking behaviors in the African setting, higher neutrophils count, lower lymphocytes, and platelet count could explain the observed findings [23, 39, 40].

Being exposed during waves 2 & 3 was a significant predictor of mortality. The wave exposure happened at different times across the world. Therefore the findings of these waves are not consistent at all compared to other settings as they occurred at different times and settings. Low mortality was observed among patients exposed to waves 1 and 4 respectively.

Different studies reported a higher mortality risk among elderly patients with one or more than two pre-existing comorbidities, especially hypertension, diabetes, and renal disease [22, 24, 31, 34, 43, 44], which was similar to the findings in this study. These studies found diabetes and hypertension as important risk factors for COVID-19 severity and mortality. The observed difference may be due to lifestyle, eating habits, sedentary lifestyle and low prevalence of Non-Communicable diseases in sub-Saharan Africa.

## Strengths

This study is the first to determine the predictors of mortality among COVID-19 confirmed cases at KCMC zonal referral hospital in Northern Tanzania. Secondly, the survival analysis which take into account variable time was carried out which make this study unique compared to others done in Tanzania. Thirdly, Data analysis and experience set a baseline for further studies in addressing the area of improvement within our country.

## Limitations

We were unable to obtain all important data related to the outcome of interest as the use of secondary data limit the prediction of the existing data and patient monitoring. Secondly, this study is single centered thus these results cannot be generalized to all COVID-19 patients admitted in the other settings. Thirdly, the external validation was limited as the studied number of patients were not sufficient enough to identify both potential predictors associated with the outcome. Fourthly, being historical cohort study, retrieving complete information was difficulty lack of some information’s might underestimate or overestimate the outcome. Lastly, Patients in this study were only studied up to their date of discharge, it would be of more interest and benefit if they were studied beyond this time. Furthermore, it also includes the usual limitations of retrospective studies.

## Conclusion

Mortality was higher in elderly male patients, with a severe form of COVID-19, who were exposed during waves 2 & 3, and those with any comorbidities. Therefore, more attention should be provided to older patients, and ensuring standard and supportive care at primary health facilities are available.

## Recommendations

Ensure that standard and supportive care are also available at the primary health facilities to allow early intervention to take place among patients diagnosed with the severe form of the disease because some patients were transferred late to referral hospitals which turnout in poor clinical outcome.

A further large multicenter quantitative study should be carried out by using both signs, symptoms, pre-existing comorbidities, laboratory parameters, and radiological and pathological findings to identify the potential predictors of mortality among COVID-19 patients, to allow external validation. Furthermore, a clinical trial study is needed to explore the sex differences in susceptibility, severity and outcomes of coronavirus diseases in 2019.

**Figure 1:**
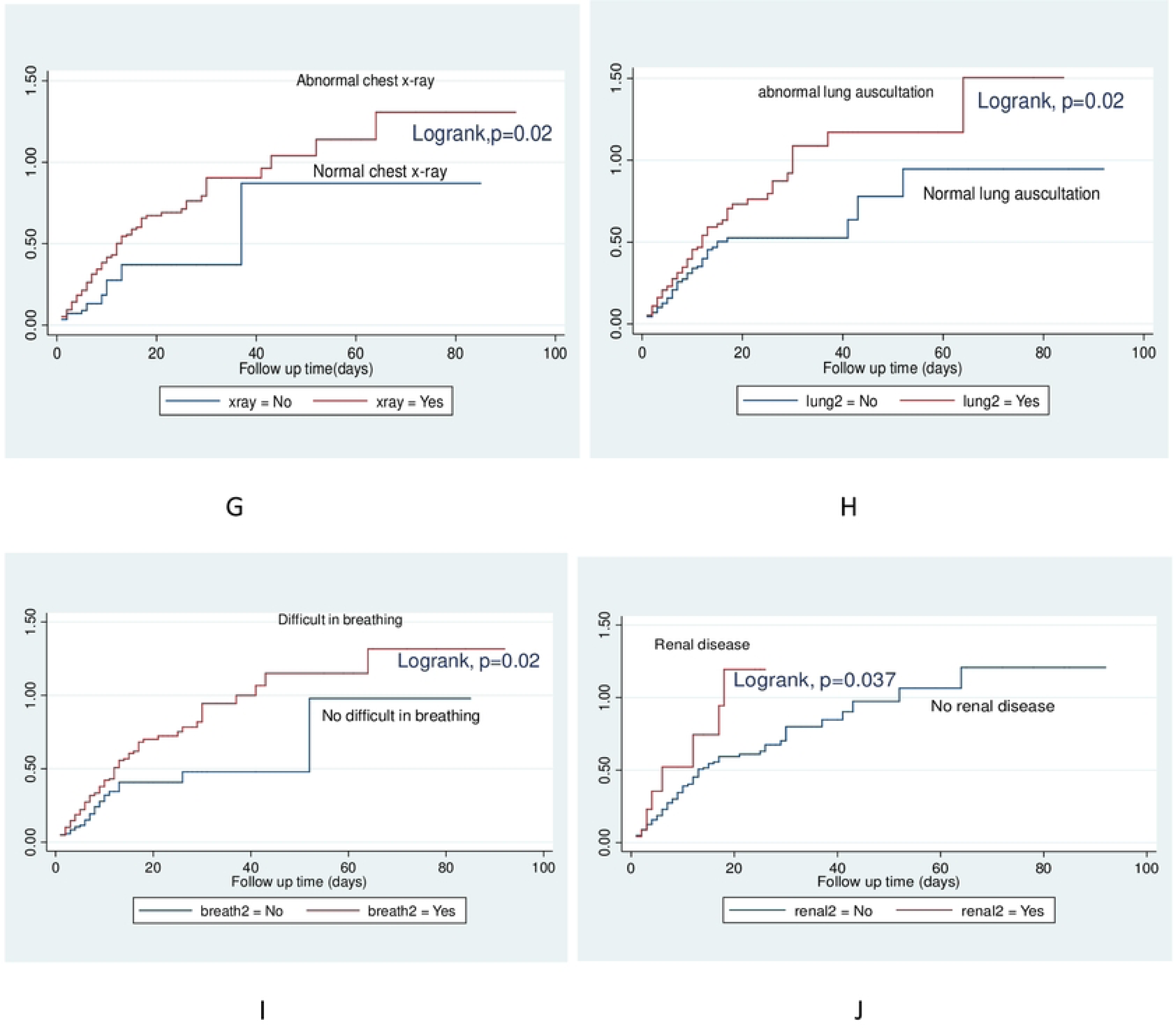
(A)Kaplan-Meier survival plot showing overall survival; and Nelson-Aalen cumulative hazard plots showing mortality experience of COVID-19 patients at KCMC Hospital from 20^th^ March 2020 to 26^th^ Feb 2022 by (B)Age; (C)Disease severity; (D)Diabetes; (E)Sex; (F)Hypertension; (G)Lung auscultations; (H)Chest-x-ray; (I)Difficult in breathing; (J)Renal disease.

## Data Availability

Data cannot be shared publicly because of restrictions of the institution. Data are available from the Kilimanjaro Christian Medical Center, Institutional Data Access / Ethics Committee (contact via +255752337067) for researchers who meet the criteria for access to confidential data.

## Abbreviations

COVID-19: Coronavirus disease of 2019
DIB: Difficulty in breathing
DM: Diabetes mellitus
GBW: Generalized Body weakness
HIV: Human Immunodeficiency Virus
IPC: Infection Prevention and Control
KCMC: Kilimanjaro Christian Medical Centre
KCMUCo: Kilimanjaro Christian Medical University College
MoH: Ministry of Health
PCR: Polymerase Chain Reaction
SARS-CoV-2: Severe Acute Respiratory Syndrome Coronavirus-2
WHO: World Health Organization.

## Consent for publication

Not applicable

## Availability of data and materials

Data will be made available

## Acknowledgment

We acknowledge all doctors at KCMC who took part in data collection and all patients whose information enabled the availability of data used in this study. The authors also thank the KCMC isolation team for capturing these data in the electronic system.

## Author’s contributions

GGW, FJM, IBM, KGK and NJ contributed to the design of the study. GGW performed the statistical analysis and drafted the manuscript. LS, HLM, EBM and MN participated in statistical analysis and drafting of the manuscript. FJM, IBM, KGK and FS edited the final draft of the manuscript and approved the final manuscript.

---

**S1 Table:**
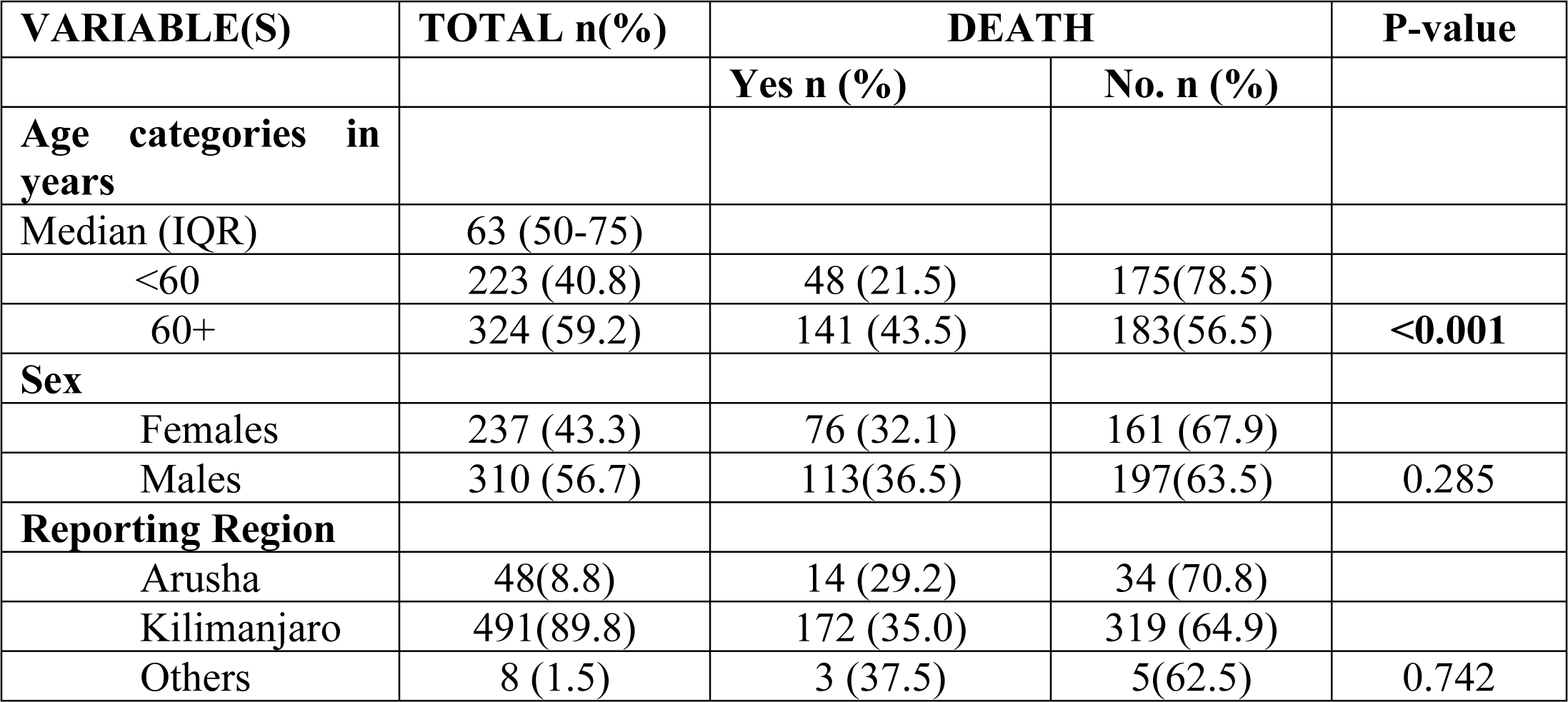
Proportions of COVID-19 patients who died by their background characteristics (N=547)

| VARIABLE(S) | TOTAL n(%) | DEATH |  | P-value |
| --- | --- | --- | --- | --- |
|  |  | Yes n (%) | No. n (%) |  |
| **Age categories in years** |  |  |  |  |
| Median (IQR) | 63 (50-75) |  |  |  |
| <60 | 223 (40.8) | 48 (21.5) | 175(78.5) |  |
| 60+ | 324 (59.2) | 141 (43.5) | 183(56.5) | **<0.001** |
| **Sex** |  |  |  |  |
| Females | 237 (43.3) | 76 (32.1) | 161 (67.9) |  |
| Males | 310 (56.7) | 113(36.5) | 197(63.5) | 0.285 |
| **Reporting Region** |  |  |  |  |
| Arusha | 48(8.8) | 14 (29.2) | 34 (70.8) |  |
| Kilimanjaro | 491(89.8) | 172 (35.0) | 319 (64.9) |  |
| Others | 8 (1.5) | 3 (37.5) | 5(62.5) | 0.742 |

**S2 Table:** Proportions of COVID-19 patients who died by clinical features at admission (N=547)

| Variables | N (%) | Died |  | P-value |
| --- | --- | --- | --- | --- |
|  |  | Yes(%) | No(%) |  |
| **Median (IQR) Oxygen saturation at admission in %** | 70(53-82) |  |  | **<0.001** |
| **Median Survival time (IQR) in days** | 7(3-12) |  |  | **0.0004** |
| **Disease severity** |  |  |  |  |
| Mild | 129 (23.6) | 17 (13.2) | 112 (86.8) |  |
| Moderate | 175 (31.9) | 32 (18.3) | 143 (81.7) |  |
| Severe | 243 (44.4) | 140 (57.6) | 103 (42.4) | **<0.001** |
| *Clinical features* |  |  |  |  |
| **Fever chills** |  |  |  |  |
| No | 214 (39.1) | 71 (33.2) | 143(66.8) |  |
| Yes | 333 (60.8) | 118 (35.4) | 215 (64.6) | 0.588 |
| **Headache** |  |  |  |  |
| No | 455 (83.2) | 159 (34.9) | 296 (65.1) |  |
| Yes | 92 (16.8) | 30 (32.6) | 62 (67.4) | 0.667 |
| **Nausea/Vomiting** |  |  |  |  |
| No | 484 (88.5) | 170 (35.1) | 314 (64.9) |  |
| Yes | 63 (11.5) | 19 (30.2) | 44 (69.8) | 0.436 |
| **Cough** |  |  |  |  |
| No | 123 (22.5) | 37 (30.1) | 86 (69.9) |  |
| Yes | 424 (77.5) | 152 (35.9) | 272 (64.1) | 0.236 |
| **Difficulty in breathing** |  |  |  |  |
| No | 146 (26.7) | 35 (23.9) | 111(76.0) |  |
| Yes | 401 (73.3) | 154 (38.4) | 247 (61.6) | **0.002** |
| **Chest pain** |  |  |  |  |
| No | 295 (53.9) | 96 (32.5) | 199 (67.5) |  |
| Yes | 252 (46.1) | 93 (36.9) | 159 (63.1) | 0.285 |
| **GBW** |  |  |  |  |
| No | 157 (28.7) | 49 (31.2) | 108 (68.8) |  |
| Yes | 390 (71.3) | 140 (35.9) | 250 (64.1) | 0.297 |
| **Sore throat** |  |  |  |  |
| No | 507 (92.7) | 179 (35.3) | 328 (64.7) |  |
| Yes | 40 (7.3) | 10 (25.0) | 30 (75.0) | 0.187 |
| **Runny nose** |  |  |  |  |
| No | 519 (94.9) | 183 (35.3) | 336 (64.7) |  |
| Yes | 28 (5.1) | 6 (21.4) | 22 (78.6) | 0.134 |
| **Smell loss** |  |  |  |  |
| No | 502 (91.7) | 178 (35.5) | 324 (64.5) |  |
| Yes | 45 (8.2) | 11 (24.4) | 34 (75.6) | 0.137 |
| **Loss of taste** |  |  |  |  |
| No | 473 (86.5) | 170 (35.9) | 303 (64.1) |  |
| Yes | 74 (13.5) | 19 (25.7) | 55 (74.3) | 0.084 |
| **Abdominal pain** |  |  |  |  |
| No | 503 (91.9) | 174 (34.6) | 329 (65.4) |  |
| Yes | 44 (8.0) | 15 (34.1) | 29 (65.9) | 0.947 |
| **Diarrhea** |  |  |  |  |
| No | 503 (91.9) | 179 (35.6) | 324 (64.4) |  |
| Yes | 44 (8.0) | 10 (22.7) | 34 (77.3) | 0.085 |
| **Waves** |  |  |  |  |
| Wave1 | 101 (18.5) | 25 (24.8) | 76 (75.3) |  |
| Wave2&3 | 375(68.6) | 133 (35.5) | 242 (64.5) |  |
| Wave4 | 71 (12.9) | 31 (43.7) | 40 (56.3) | **0.030** |

**S3 Table:**
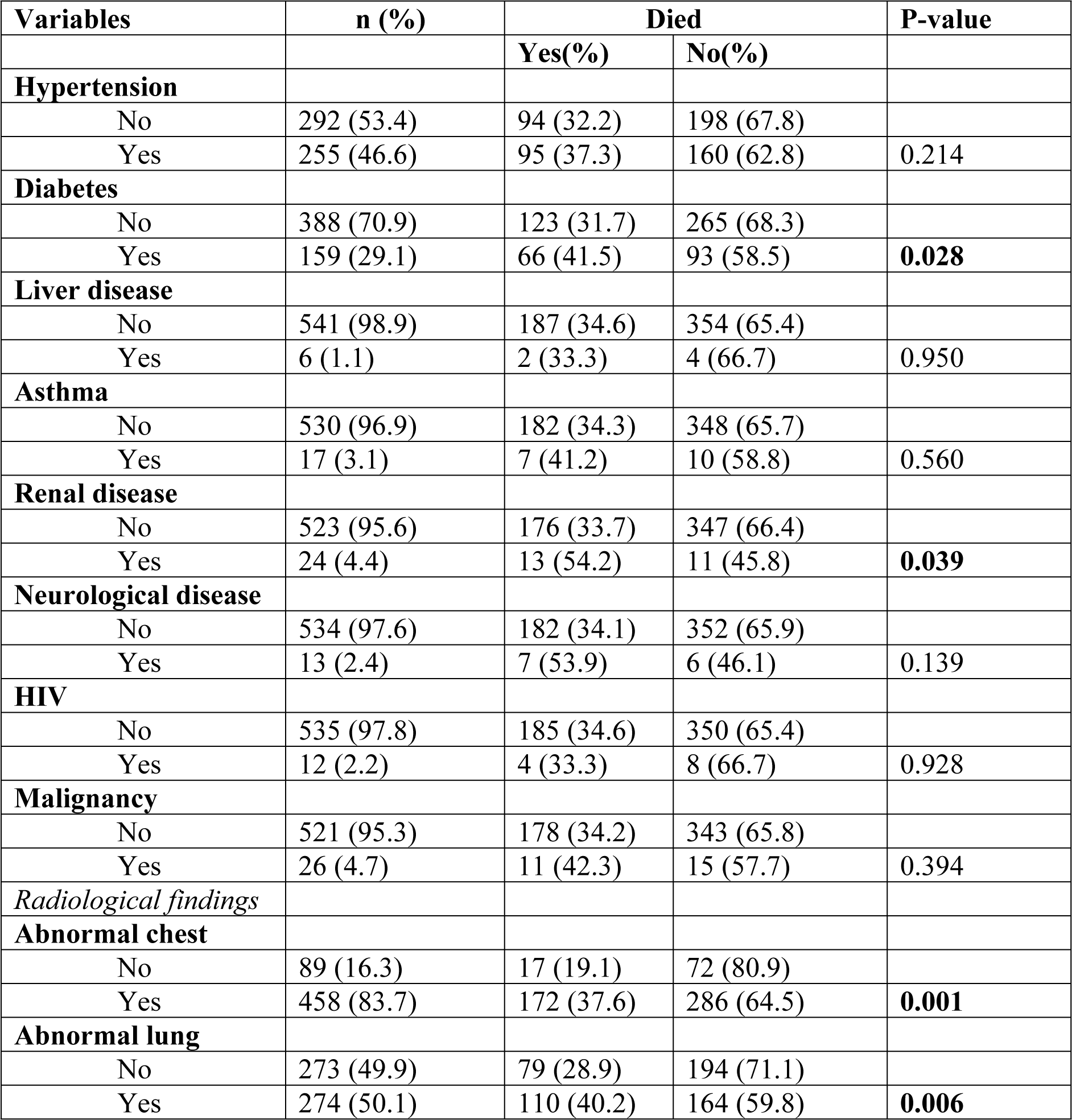
Proportion of COVID-19 patients who died by comorbidities and radiological findings (N=547)

**S4 Table:**
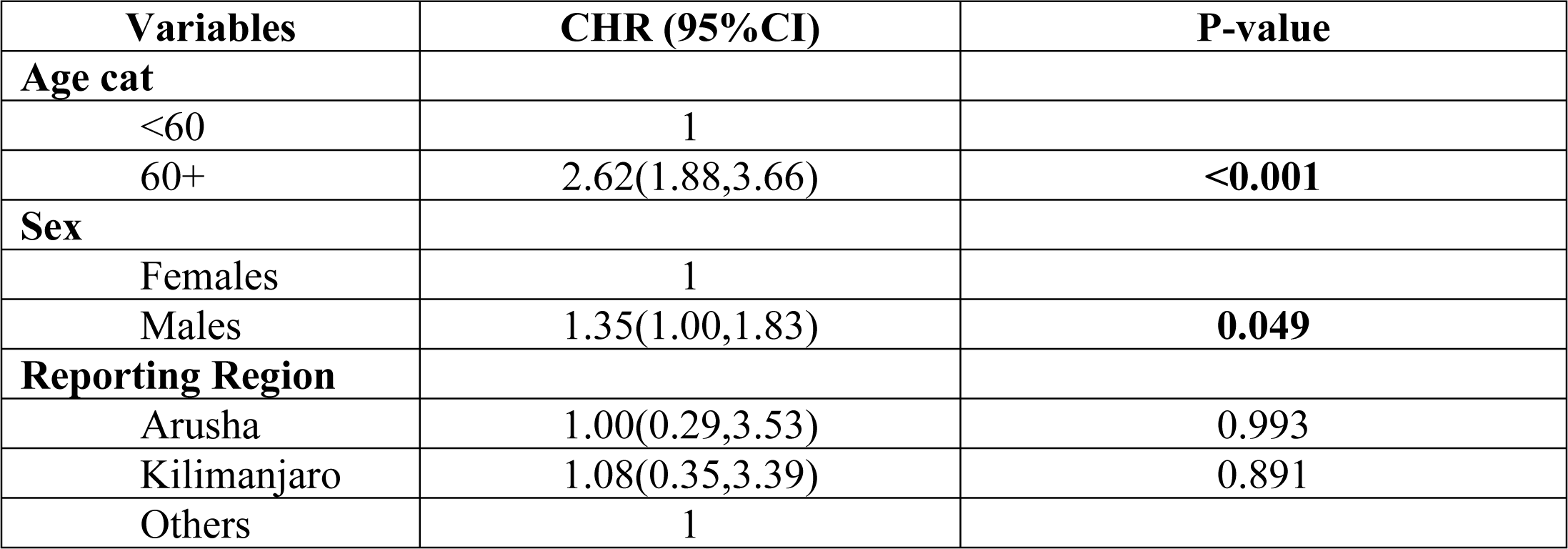
Sociodemographic predictors of mortality among COVID-19 patients.

| **Variables** | **CHR (95%CI)** | **P-value** |
| --- | --- | --- |
| **Age cat** |  |  |
| <60 | 1 |  |
| 60+ | 2.62(1.88,3.66) | **<0.001** |
| **Sex** |  |  |
| Females | 1 |  |
| Males | 1.35(1.00,1.83) | **0.049** |
| **Reporting Region** |  |  |
| Arusha | 1.00(0.29,3.53) | 0.993 |
| Kilimanjaro | 1.08(0.35,3.39) | 0.891 |
| Others | 1 |  |

**S5 Table:**
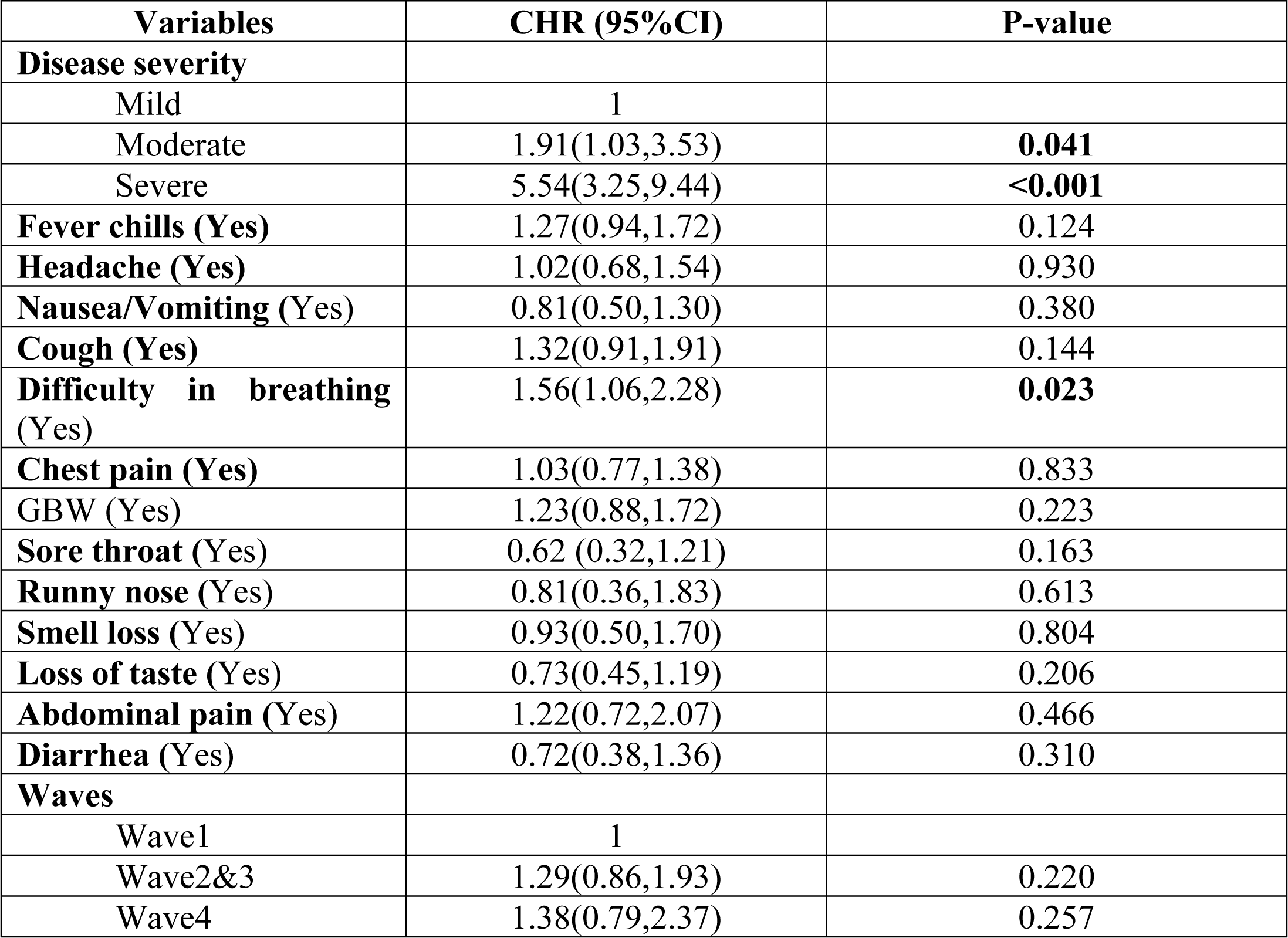
Clinical features predictors associated with mortality among COVID-19 patients.

**S6 Table:** Association between comorbidities, radiological findings and COVID-19 mortality.

| **VARIABLE(S)** | **CHR (95%CI)** | **P-value** |
| --- | --- | --- |
| **Hypertension** |  |  |
| Yes | 1.27(0.95,1.71) | 0.109 |
| **Diabetes** |  |  |
| Yes | 1.33(0.98,1.82) | **0.066** |
| **Liver disease** |  |  |
| Yes | 1.00(0.25,4.05) | 0.995 |
| **Asthma** |  |  |
| Yes | 0.83(0.39,1.79) | 0.652 |
| **Renal disease** |  |  |
| Yes | 1.89(1.08,3.33) | **0.027** |
| **Neurological disease** |  |  |
| Yes | 1.87(0.87,3.97) | 0.106 |
| **Immunodeficiency** |  |  |
| Yes | 0.54(0.20,1.47) | 0.228 |
| **Malignancy** |  |  |
| Yes | 1.18(0.64,2.17) | 0.596 |
| *Radiological findings* |  |  |
| **Abnormal chest x-ray** |  |  |
| Yes | 1.75(1.06,2.88) | **0.028** |
| **Abnormal lung auscultation** |  |  |
| Yes | 1.41(1.05,1.89) | **0.024** |

**S7 Table:**
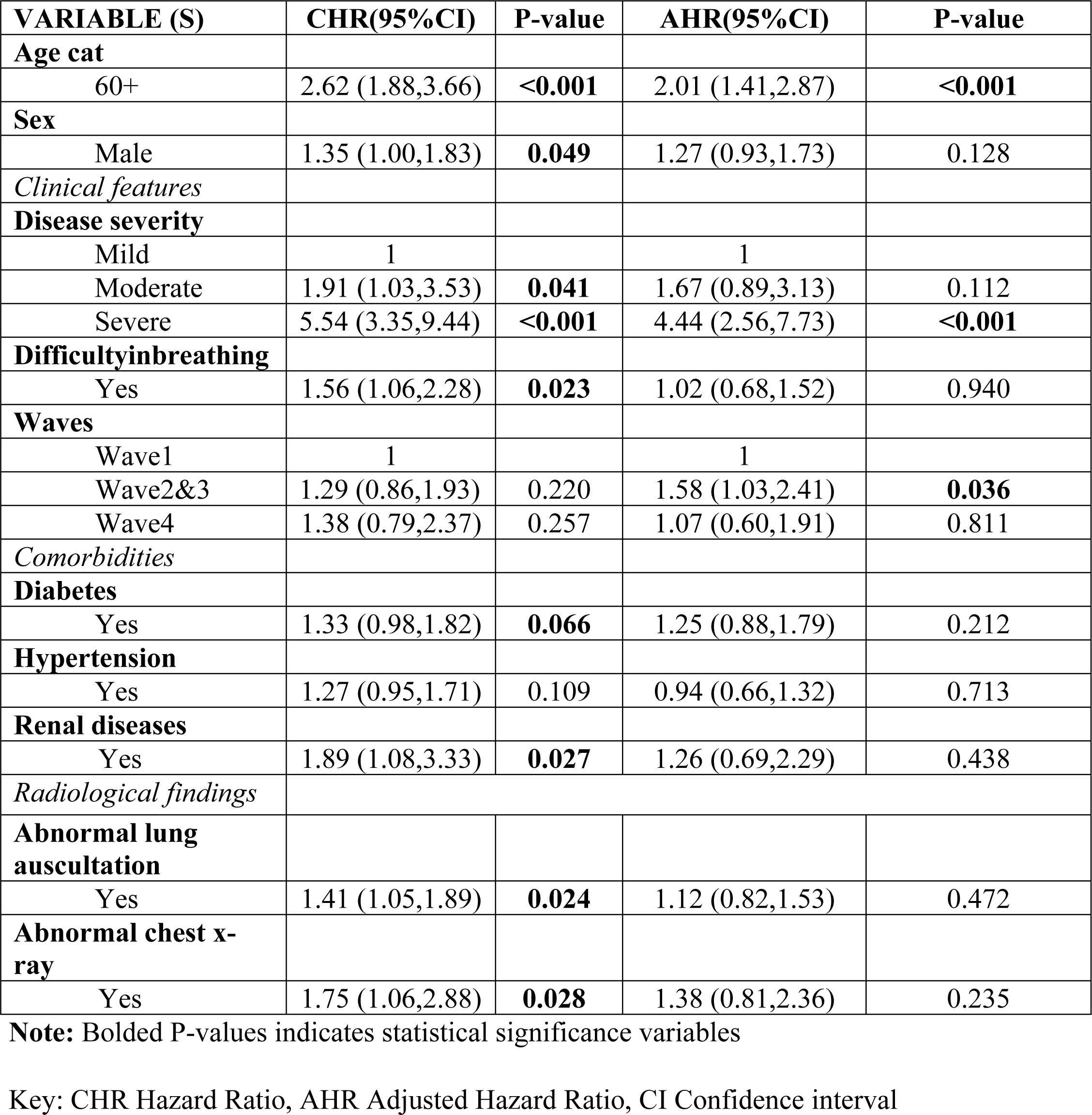
Predictors of mortality among COVID-19 patients.

| **VARIABLE (S)** | **CHR(95%CI)** | **P-value** | **AHR(95%CI)** | **P-value** |
| --- | --- | --- | --- | --- |
| **Age cat** |  |  |  |  |
| 60+ | 2.62 (1.88,3.66) | **<0.001** | 2.01 (1.41,2.87) | **<0.001** |
| **Sex** |  |  |  |  |
| Male | 1.35 (1.00,1.83) | **0.049** | 1.27 (0.93,1.73) | 0.128 |
| *Clinical features* |  |  |  |  |
| **Disease severity** |  |  |  |  |
| Mild | 1 |  | 1 |  |
| Moderate | 1.91 (1.03,3.53) | **0.041** | 1.67 (0.89,3.13) | 0.112 |
| Severe | 5.54 (3.35,9.44) | **<0.001** | 4.44 (2.56,7.73) | **<0.001** |
| **Difficultyinbreathing** |  |  |  |  |
| Yes | 1.56 (1.06,2.28) | **0.023** | 1.02 (0.68,1.52) | 0.940 |
| **Waves** |  |  |  |  |
| Wave1 | 1 |  | 1 |  |
| Wave2&3 | 1.29 (0.86,1.93) | 0.220 | 1.58 (1.03,2.41) | **0.036** |
| Wave4 | 1.38 (0.79,2.37) | 0.257 | 1.07 (0.60,1.91) | 0.811 |
| *Comorbidities* |  |  |  |  |
| **Diabetes** |  |  |  |  |
| Yes | 1.33 (0.98,1.82) | **0.066** | 1.25 (0.88,1.79) | 0.212 |
| **Hypertension** |  |  |  |  |
| Yes | 1.27 (0.95,1.71) | 0.109 | 0.94 (0.66,1.32) | 0.713 |
| **Renal diseases** |  |  |  |  |
| Yes | 1.89 (1.08,3.33) | **0.027** | 1.26 (0.69,2.29) | 0.438 |
| *Radiological findings* |  |  |  |  |
| **Abnormal lung auscultation** |  |  |  |  |
| Yes | 1.41 (1.05,1.89) | **0.024** | 1.12 (0.82,1.53) | 0.472 |
| **Abnormal chest x-ray** |  |  |  |  |
| Yes | 1.75 (1.06,2.88) | **0.028** | 1.38 (0.81,2.36) | 0.235 |

**Note:** Bolded P-values indicates statistical significance variables
Key: CHR Hazard Ratio, AHR Adjusted Hazard Ratio, CI Confidence interval

## REFERENCES

1. WHO (2020) Coronavirus Disease (COVID-19) Situation Reports Updates 27 September 2020. World Health Organ Tech Rep Ser 1–23

2. WHO (2022) COVID-19 Weekly Epidemiological Update. World Heal Organ 1–23

3. Spiteri G, Fielding J, Diercke M, et al (2020) First cases of coronavirus disease 2019 (COVID-19) in the WHO European Region, 24 January to 21 February 2020. 2019:

4. Alshogran OY, Altawalbeh SM, Al-azzam SI, Karasneh R (2021) Predictors of Covid-19 case fatality rate: An ecological study. Ann Med Surg 65:102319

5. Ferroni E, Rossi PG, Alegiani SS, et al (2020) Survival of hospitalized COVID-19 patients in Northern Italy: A population-based cohort study by the ITA-COVID-19 network. Int J Environ Res Public Health 17:1–12

6. Lei J, Li M, Wang X (2021) Predicting the development trend of the second wave of COVID-19 in five European countries. J Med Virol 93:5896–5907

7. Wang Y, Hospital CF, Ren L, Fan G, Hospital CF, Gu X, Hospital CF (2020) Articles Clinical features of patients infected with 2019 novel coronavirus in Wuhan, China. 10.1016/S0140-6736(20)30183-5

8. Emami A, Javanmardi F, Akbari A, Kojuri J, Bakhtiari H, Rezaei T, Keshavarzi A, Falahati F (2021) Survival rate in hypertensive patients with. Clin Exp Hypertens 43:77–80

9. Lot M, Hamblin MR, Rezaei N (2020) Since January 2020 Elsevier has created a COVID-19 resource centre with free information in English and Mandarin on the novel coronavirus COVID-19. The COVID-19 resource centre is hosted on Elsevier Connect, the company’s public news and information.

10. Mohammed M, Muhammad S, Mohammed FZ, Mustapha S, Sha A, Sani NY, Ahmad MH, Bala AA (2020) Risk Factors Associated with Mortality Among Patients with Novel Coronavirus Disease (COVID-19) in Africa. 6–11

11. Rosenthal PJ, Breman JG, Djimde AA, John CC, Kamya MR, Leke RGF, Moeti MR, Nkengasong J, Bausch DG (2020) Editorial COVID-19: Shining the Light on Africa. 102:1145–1148

12. Medhat MA, Kassas M El (2020) COVID-19 in Egypt: Uncovered figures or a different situation? 10:2–5

13. Sarvas J (1987) Basic mathematical and electromagnetic concepts of the biomagnetic inverse problem. Phys Med Biol 32:11–22

14. MoHCDGEC (2021) THE UNITED REPUBLIC OF TANZANIA Weekly trend of COVID-19 Confirmed Cases. 1–7

15. Goyal ADK, Mansab AF, C Bai, D sb (2020) Early intervention likely improves mortality in COVID-19 infection. 248–250

16. Nlandu Y, Mafuta D, Sakaji J, et al (2021) Predictors of mortality in COVID - 19 patients at Kinshasa Medical Center and a survival analysis: a retrospective cohort study. BMC Infect Dis 1–11

17. Methods C, Vienna LS, Vienna LS, Ducrocq V (2013) The Survival Kit: Software to analyze survival data including possibly correlated random effects. 10.1016/j.cmpb.2013.01.010

18. Aghan E, Aziz O, Mbithe H, Surani S, Orwa J, Somji S (2021) Factors Associated with Mortality Among Hospitalized Adults with COVID-19 Pneumonia at a Private Tertiary Hospital in Tanzania: A Retrospective Cohort Study. 5431–5440

19. Mammen JJ, Kumar S, Thomas L, et al (2021) Factors associated with mortality among moderate and severe patients with 19 in India: a secondary of a randomised controlled trial. 1–10

20. Mnyambwa NP, Lubinza C, Ngadaya E, et al (2022) Clinical characteristics and outcomes of confirmed COVID-19 patients in the early months of the pandemic in Tanzania: a multicenter cohort study. IJID Reg 2:118–125

21. Agarwal N, Biswas B, Singh C, Nair R, Mounica G, Haripriya H, Jha AR, Das KM (2021) Early Determinants of Length of Hospital Stay: A Case Control Survival Analysis among COVID-19 Patients admitted in a Tertiary Healthcare Facility of East India. 10.1177/21501327211054281

22. Weya A, Id K, Id GA, Hurissa Z, Kaso T, Ewune HA, Hareru HE, Id AH (2022) Survival analysis of COVID-19 patients in Ethiopia: A hospital-based study. 1–11

23. Lackcy M c. B and elisabeth m. (2020) Categorization of COVID-19 severity to determine mortality risk. FDA 19

24. Ayana galana mamo (2021) Predictors of Mortality Among Hospitalized COVID-19 Patients at a Tertiary Care Hospital in. Infect drug Resist 5363–5373

25. Pereira MLD (2020) Mortality and survival of COVID-19. Epidemilogy Infect 148:1– 6

26. Ciceri F, Castagna A, Rovere-querini P, et al (2020) Early predictors of clinical outcomes of COVID-19 outbreak in Milan, Italy. Clin Immunol 217:108509

27. Bwire GM (2020) Coronavirus: Why Men are More Vulnerable to Covid-19 Than Women? 874–876

28. Wahaibi (2020) COVID-19 disease severity and mortality determinants: A large population-based analysis in Oman. Travel Med. Infect. Dis. 39 101923 Contents

